# Association of lung clearance index with survival in individuals with cystic fibrosis

**DOI:** 10.1101/2021.06.25.21254759

**Authors:** Johanna Manuela Kurz, Kathryn Angela Ramsey, Romy Rodriguez, Ben Spycher, Reta Fischer Biner, Philipp Latzin, Florian Singer

**Affiliations:** Division of Respiratory Medicine, Department of Paediatrics, Inselspital University Hospital Bern, University of Bern, Bern, Freiburgstrasse 15, 3010 Bern, Switzerland; Graduate School for Health Sciences, University of Bern, Bern, Switzerland; Institute of Social and Preventive Medicine, University of Bern, Mittelstrasse 43, 3012 Bern, Switzerland; Quartier Bleu, Lindenhofspital, Bremgartenstrasse 117, 3012 Bern, Switzerland

**Keywords:** Respiratory function tests, Cystic fibrosis, Analysis, Survival, Mortality, Lung transplantaion

## Abstract

**Background:** Lung clearance index (LCI) quantifies global ventilation inhomogeneity, a sensitive biomarker of airway function in cystic fibrosis (CF) lung disease.

**Objectives:** We examined the association of LCI with the risk of death or lung transplantation (LTX) in individuals with CF.

**Methods:** We performed a retrospective analysis in a cohort of individuals with CF aged ≥ five years with available LCI and FEV_1_ measurements between 1980 and 2006. Outcome was time until death or LTX. We applied Cox proportional hazard regressions using the earliest available LCI and FEV_1_ values and adjusted for demographic and clinical variables. For sensitivity analyses, we used the mean of the first three LCI and FEV_1_ measurements, stratified the cohort based on age, and investigated individuals with normal FEV_1_.

**Results:** In total, 237 individuals with CF aged mean (range) 13.9 (5.6–41.0) years were included. This time-to-event analysis accrued 3813 person-years, 94 (40%) individuals died or received LTX. Crude hazard ratios [95% CI] were 1.04 [1.01–1.06] per one z-score increase in LCI and 1.25 [1.11–1.41] per one z-score decrease in FEV_1_. After adjusting LCI and FEV_1_ mutually in addition to sex, age, BMI and the number of hospitalisations, hazard ratios were 1.04 [1.01-1.07] for LCI, and 1.12 [0.95-1.33] for FEV_1_. Sensitivity analyses yielded similar results and using the mean LCI strengthened the associations.

**Conclusions:** Increased ventilation inhomogeneity is associated with greater risk of death or LTX. Our data support LCI as novel surrogate of survival in individuals with CF.

**TAKE HOME MESSAGE:** Lung clearance index (LCI) is a measure of global ventilation inhomogeneity which increases early during the course of Cystic Fibrosis (CF) lung disease. This study shows that LCI is predictive of death or lung transplantation in individuals with CF.

**Study registration number:** NCT04016194

## Introduction

Cystic fibrosis (CF) is the most prevalent inherited lethal multi-organ disease in the general Caucasian population[1]. The progression of chronic lung disease leads to lung function decline and respiratory failure, which remains the major cause of morbidity and mortality[2]. Spirometry derived forced expired volume in the first second (FEV_1_) is used as physiological surrogate to predict survival and for referral to lung transplantation (LTX)[3]. Within the past 20 years, the progression of CF lung disease has slowed and FEV_1_ is often normal despite physiological and radiological signs of subclinical lung disease[4]. In individuals with mild CF lung disease, FEV_1_ is less responsive to treatment[5]. The lung clearance index (LCI) derived from multiple breath inert gas washout (MBW) quantifies global ventilation inhomogeneity and is a sensitive biomarker of central and peripheral airway function[6]. MBW is safe and requires minimal patient cooperation[7]. LCI is more strongly correlated with structural lung damage than FEV_1_[4]. However, the clinical interpretation of increased (abnormal) LCI is currently constrained, as the ability to predict adverse respiratory disease outcomes has not been established yet[8]. In particular, it remains unclear if LCI can be considered as a surrogate endpoint for survival.

We utilised an existing cohort of individuals with CF, who were longitudinally followed including MBW measurements performed during routine clinical visits since the 1980s[9]. We hypothesized that individuals with CF who have elevated LCI are at increased risk of death or LTX. The objective of this study was to determine the association of LCI with adverse respiratory disease outcomes. The secondary aim was to compare the predictive values of the two parameters LCI and FEV_1_.

## Methods

### Study design

We performed a retrospective, observational analysis (NCT04016194) in a cohort of paediatric and adult individuals with CF at the paediatric CF outpatient clinic at the University Children’s Hospital Bern, Switzerland. Source data were electronic patient records. We collated lung function and clinical data within three years after the first available MBW measurement. Routine assessments at each clinical visit were: MBW measurements using nitrogen (N_2_) as tracer gas (N_2_MBW) and spirometry, microbiological culture from cough swabs or sputum, body height and weight, records of medication, number of exacerbations, and hospitalisations[10]. Lung function indices from spirometry but not MBW were routinely disclosed to treating clinicians. Ethical approval was obtained by the local ethics committee (KEK BE 2018-01642). Written informed consent from survivors and relatives of the individuals who died was not required from the ethics committee and therefore not obtained for this study.

### Inclusion- and exclusion criteria

Eligible individuals were screened by systematic review. Eligibility criteria were: Confirmed CF diagnosis, age ≥ five years, available records on routine clinical care in the paediatric CF-centre Bern. Inclusion criteria were: Availability of at least one N_2_MBW and FEV_1_ measurement between 1986/01/01-2006/12/31. At that time, CF was diagnosed clinically in presence of typical respiratory symptoms or failure to thrive, two disease-causing CFTR mutations, and abnormal sweat chloride[11]. We excluded individuals if CF diagnosis was not confirmed, or if lung function tests were available only after LTX.

### Variables and definitions

Outcome was defined as time until death or LTX. End of study was on 2018/12/31. We refer to survival as respiratory survival, where death and LTX are regarded as equivalent markers for terminal pulmonary disease as previously described[12]. We included LCI as primary and FEV_1_ as secondary predictor variable. We *a priori* selected the following variables to account for possible confounding[13]: year of birth, age at CF diagnosis, age at lung function measurement, sex, and body mass index (BMI, normalized to z-scores[14]). Birth year was used as proxy for temporal trends in medical care. In addition, the following clinical variables were investigated[15-17]: genotype, pancreas function, CF-related diabetes (CFRD), infection with *Staphylococcus aureus* and *Pseudomonas aeruginosa*, allergic bronchopulmonary aspergillosis (ABPA), antibiotic treatment, and the number of pulmonary exacerbations, and hospitalisations. All variables collected from the source data were entered into an online database (RedCap database)[18]. Variable definitions are summarized in Table S1, online supplement (OLS). Age was used as the underlying analysis time. Children entered the study at the age of the first LCI measurement and contributed time at risk until the event (death or LTX), loss to follow-up or the end of the study period, whichever occurred first. Time at risk of individuals who were lost to follow-up was right censored on the date of the last available visit.

### Lung function measurement

Trained lung function technicians performed N_2_MBW according to in-house measurement standards using a customized open-bypass setup (SensorMedics 2200, Yorba Linda, CA, USA)[19]. The N_2_MBW setup and LCI analysis remained unmodified throughout the whole study period. N_2_MBW analysis was performed off-line; LCI was calculated according to the recommendations at that time: cumulative expired volume divided by function residual capacity[10, 19]. LCI values are dimensionless “lung turnovers” and referred to LCI units[7]. Generally applicable reference equations for MBW indices do not currently exist[20]. Therefore, LCI was standardized to z-scores using the distributional estimates mean (standard deviation, SD) of LCI derived from the same N_2_MBW setup in 54 healthy subjects aged between 7 and 16 years [19]: 7.64 (0.86) units. Z-scores were calculated as:

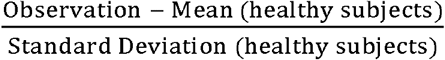

Spirometry was performed using a commercial setup (Jaeger Würzburg, Germany)[10] and in accordance with standards of the European Respiratory Society (ERS) and American Thoracic Society (ATS) at that time[21-23]. To assess lung function independent from sex, age, and height, data were expressed as z-scores[24]. The Global Lung Initiative (GLI) reference equations include data collected before 2006 and were therefore considered applicable.

We additionally expressed LCI and FEV_1_ values as standard deviation (SD-score) score based on the CF study population to account for unequal variances of LCI and FEV_1_ in the CF population:

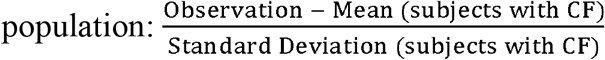

### Statistical methods

In the main analysis, we fitted Cox proportional hazard regressions to investigate the association of baseline LCI with survival *vs*. death or LTX with age (years) as the underlying time variable. The baseline was defined as the first available LCI value (date of study entry), combined with the clinical information derived within the three subsequent years after study entry. In sensitivity analyses, we first included only individuals with at least three available LCI values and used the mean of the first three available LCI and FEV_1_ measurements within three years as baseline values. Average lung function values were expected to account for variability in lung function values between visits and to avoid possible confounding by indication[25]. As none of the included individuals received LTX or died within the first three years, the prediction started at the date of the third available LCI value and immortal time bias was avoided. Second, we stratified individuals based on age and repeated the analysis using the initial baseline definition (first available LCI and FEV_1_ measurement) as in the main analysis (i) in children aged ≤ 16.0 years at baseline, (ii) in individuals born within 30 years prior study end (excluding individuals born earlier than 1987) and (iii) in adults aged > 16.0 years at baseline. Third, we investigated the association of LCI with death or LTX including only individuals with normal FEV_1_ (FEV_1_ ≥ -1.96 z-score). We report unadjusted and adjusted estimates (Hazard Ratios, HR), and 95% confidence intervals (CI). The Cox proportional hazard regression analysis was performed in five steps (Figure S1, OLS): *(i) Crude model:* unadjusted, including LCI and FEV_1_ separately; *(ii) Mutual model:* including both, LCI and FEV_1_; *(iii) Complete model:* including LCI and FEV_1_ separately, adjusted for all demographic and clinical variables; *(iv) Reduced model:* including LCI and FEV_1_ separately, adjusted for selected demographic and clinical variables only; *(v) Final model:* including both, LCI and FEV_1_, adjusted for selected demographic and clinical variables. We selected all clinical variables *a priori*, variable reduction in the reduced and final models were based on examining correlations between clinical variables and stepwise removing variables with p > 0.2 in likelihood-ratio tests.

We computed Kaplan Meier survival-curves for the following two groups: *(i)* Individuals with baseline LCI below the study population median, and *(ii)* individuals above the study population median. Analyses were performed using Stata 14.2 software package (StataCorp LP, College Station, TX, USA). P-values of <0.05 were considered statistically significant.

## Results

### Study participants

In total, 263 individuals ≥ five years of age and treated in the CF-centre Bern between 1980-2006 were assessed for eligibility (Figure 1). Lung function data was available in 237 individuals with CF. Included individuals (n = 237, 47.7% females) were born between 1952-2000 and accrued 3813.3 person-years at risk during the study period. At baseline, mean (SD) age was 13.9 (8.2) years with a range from 5.6 to 41.0 years (Table 1). Mean (SD) LCI and FEV_1_ was 8.7 (7.3) z-score and -2.5 (2.0) z-score, respectively. Average time between visits within the baseline was 10.0 (6.1) months, mean duration of follow-up was 16.1 (6.6) years. The majority of subjects (73.4%) were followed annually, 36.7% were followed semi-annually across the baseline period. Seventy individuals (39.7%) received LTX or died by 2018/12/31. Mean (SD) age at death or LTX was 30.0 (10.0) years. Fifteen individuals (6.33%) were lost to follow-up within the study period and 143 individuals (60.3%) were alive at the end of the study (right censored). Reasons for loss to follow-up were clinical care elsewhere (n = 10) and moving abroad (n = 5). Compared to individuals who received LTX, individuals who died were older but had similar FEV_1_, LCI and BMI at baseline. Follow-up duration was comparable between individuals who died and those who received LTX, further details are provided in Table S2, OLS. All included variables contained less than 5.0% of missing values.

**Table 1.** Population characteristics and study end-points.

|  | **Baseline** |
| --- | --- |
| N [females, %] | 237 [96, 51.1] |
| Year of birth | 1983 [10.7; 1952 – 2000] |
| Median age at CF diagnosis [IQR, range] | 0.0 [0.0 – 2.0; 0.0 – 28.0] |
| Age at prediction start (years) | 13.9 [8.2; 5.6 – 41.0] |
| BMI at prediction start (z-score) | -0.9 [1.1; -4.0 – 2.4] |
| F508del homozygous, n [%] | 136 [57.4] |
| F508del heterozygous, n [%] | 74 [31.2] |
| Other, n [%] | 27 [11.4] |
| Pancreatic insufficiency, n [%] | 200 [84.4] |
| CFRD, n [%] | 49 [20.7] |
| LCI (units) | 17.5 [7.3; 3.5 – 59.8] |
| LCI (z-score) | 8.7 [7.3; -5.4 – 50.9] |
| FEV<sub>1</sub> (z-score) | -2.4 [2.0; -6.4 – 1.7] |
| FEV<sub>1</sub> (%pred.) | 70.3 [24.7; 19.8 – 120.8] |
| Pseudomonas aeruginosa, n [%] | 173 [73.0] |
| Staphylococcus aureus, n [%] | 137 [57.8] |
| ABPA, n [%] | 19 [8.0] |
| Individuals hospitalized at least once during baseline, n (%) | 107 [45.2] |
| Number of hospitalisations per individual during baseline | 2.2 [1.7; 1.0 – 9.0] |
| Antibiotic treatment (oral, inhaled, intravenous), n (%) | 201 [84.1] |
| Inhaled medication (mucolytics, bronchodilators, steroids), n (%) | 204 [86.1] |
| Follow-up duration (years) until outcome event or end of study | 16.1 [6.6; 0.3 – 32.3] |
| Outcome (Death or LTX), n [%] | 94 [39.7] |
| Death, n [%] | 41 [17.3] |
| males, n [%] | 22 [53.7] |
| females, n [%] | 19 [46.3] |
| LTX, n [%] | 53 [22.4] |
| males, n [%] | 27 [50.9] |
| females, n [%] | 26 [49.1] |
| Loss to follow-up, n [%] | 15 [6.3] |
| Person-years at risk | 3813.3 |

*Definitions:* Baseline = First available LCI (= study entry) and corresponding FEV<sub>1</sub> values;
Demographic and clinical data derived within the first three years after study entry. Prediction
start = Date of third LCI value within the first three years after study entry; Follow-Up:
Duration (years) from prediction start until outcome event or end of study in 2018/12/30.
*Abbreviations:* ABPA = Allergic bronchopulmonary aspergillosis, BMI = Body mass index,
CF = Cystic fibrosis, CFRD = Cystic fibrosis-related diabetes, FEV<sub>1</sub> = Forced expired volume in the first second, LCI = Lung clearance index, LTX = Lung transplantation.

**Figure 1.**
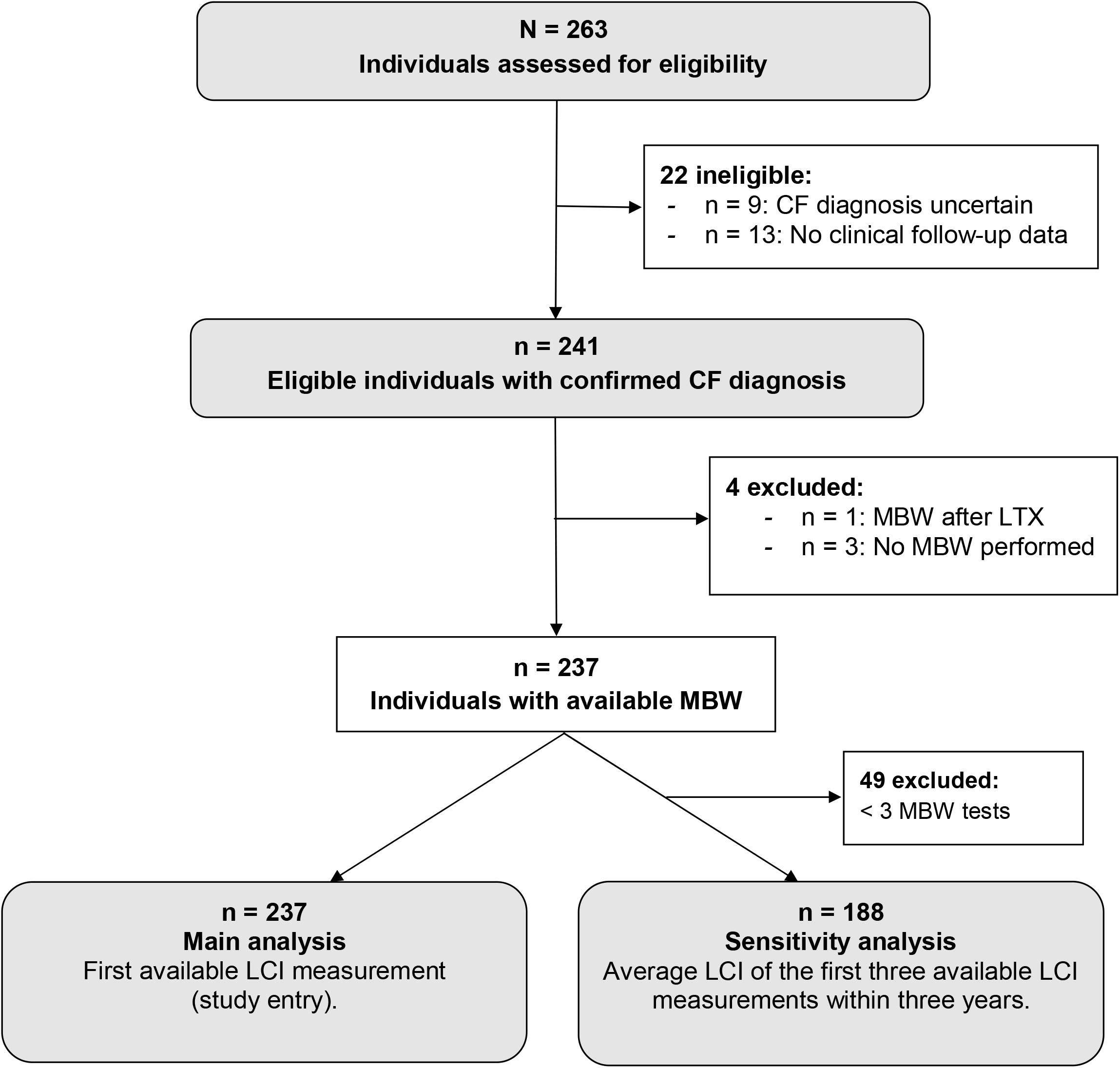
Participant flow diagram. *Eligibility criteria:* Age ≥ 5 years, confirmed CF diagnosis, routine clinical care in CF-Centre Bern between 1986–2006. *Inclusion criteria:* Availability of at least one MBW test. CF = Cystic fibrosis, LCI = Lung clearance index, MBW = Multiple breath washout, LTX = Lung transplantation.

### Main findings

Higher baseline LCI and lower baseline FEV_1_ were associated with increased risk (HR) of death or LTX in individuals with CF. In the main analysis, estimated HR [95% CI] for death or LTX from the *Crude model* were 1.04 [1.01 – 1.06] per 1.0 z-score increase in LCI and 1.25 [1.11 – 1.41] per 1.0 z-score increase in FEV_1_ (Table 2). After adjusting for selected demographic and clinical variables (sex, age, BMI, year of birth and the number of hospitalisations) in the *Reduced model* the HR was 1.04 [1.01 – 1.07] for LCI and 1.18 [1.01 – 1.38] for FEV_1_. Consequently, per 1.0 z-score increase in LCI, the risk of dying or receiving LTX increased by 4%. Per 1.0 z-score decrease in FEV_1_, the risk of dying or receiving LTX increased by 18%. Therefore, an increase by 2.4 z-score in LCI or a 0.6 z-score decrease in FEV_1_, were associated with the same risk increase of 10%. In the *Final model*, we mutually adjusted LCI and FEV_1_ in addition to the aforementioned variables. In this model, the HR for LCI was 1.04 [1.01 – 1.07]. The corresponding HR for FEV_1_ was no longer statistically significant: 1.12 [0.95 – 1.33]. Results are displayed in Figure 2. Estimates from the *Mutual* and *Complete models* can be found in Table S3, OLS. Further details on the *Complete* and *Final* models are given in Tables S6 and S7, OLS.

**Table 2.** Risk of death or lung transplantation according to baseline lung function.

| **A)** | **LCI (z-score)** | **FEV<sub>1</sub> (z-score)** |
| --- | --- | --- |
| Crude model | 1.04 [1.01 – 1.06], p = 0.006 | 1.25 [1.11 – 1.41], p < 0.001 |
| Reduced model | 1.04 [1.01 – 1.07], p = 0.003 | 1.18 [1.01 – 1.38], p = 0.043 |
| Final model | 1.04 [1.01 – 1.07], p = 0.011 | 1.12 [0.95 – 1.33], p = 0.164 |
| **B)** | **LCI (SD-score)** | **FEV<sub>1</sub> (SD-score)** |
| Crude model | 1.30 [1.08 – 1.58], p = 0.006 | 1.55 [1.22 – 1.96], p < 0.001 |
| Reduced model | 1.34 [1.10 – 1.62], p = 0.003 | 1.38 [1.01 – 1.88], p = 0.043 |
| Final model | 1.30 [1.06 – 1.60], p = 0.011 | 1.26 [0.91 – 1.74], p = 0.164 |

**Figure 2.**
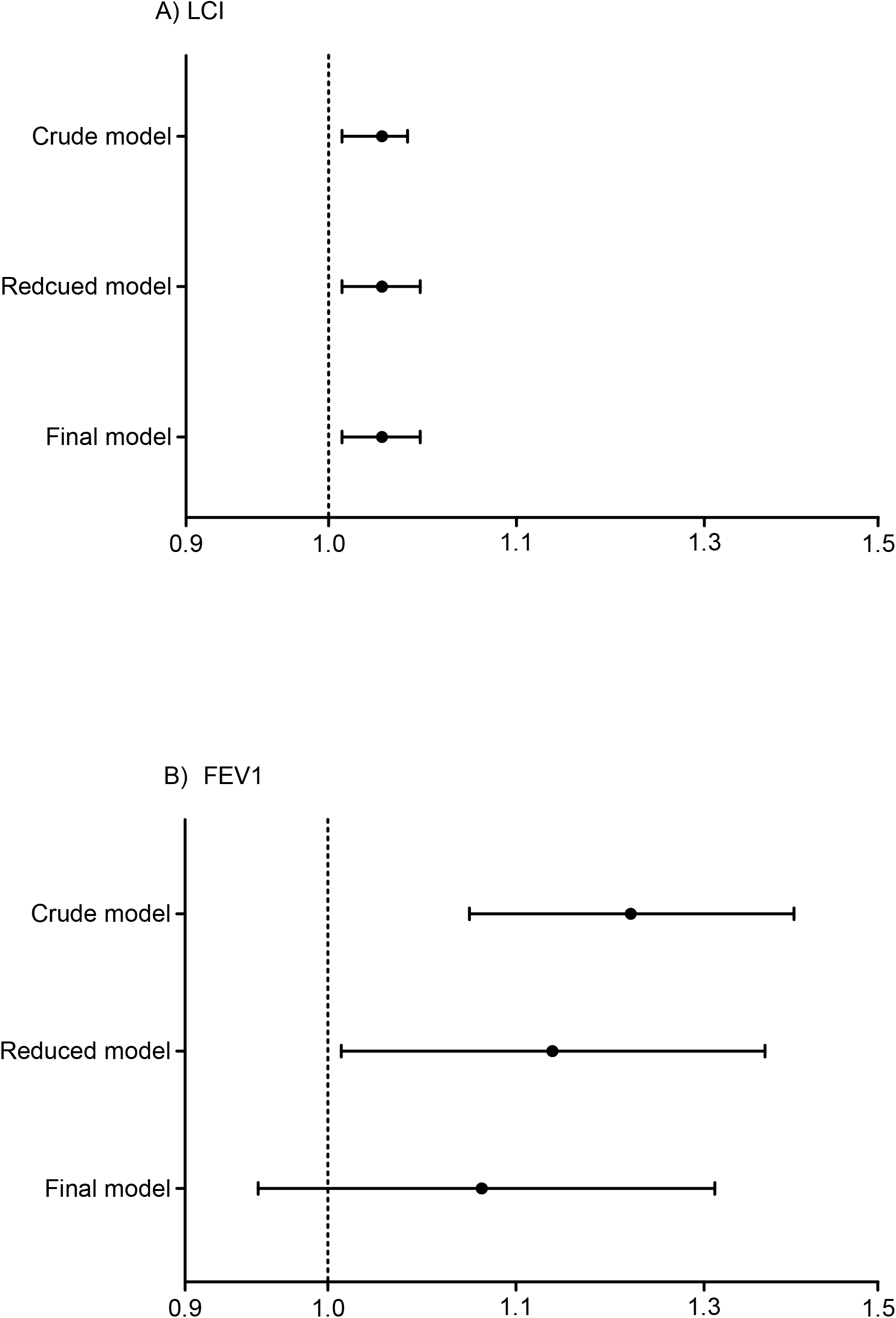
Risk of death or lung transplantation according to baseline lung function. Crude and adjusted Hazard Ratios [95% CI] for the risk of death or lung transplantation per one z-score increase in LCI (left panel) or per one z-score decrease in FEV_1_ (right panel) in 237 individuals with CF using the first available LCI and corresponding FEV_1_ value as baseline. X-axis shows log-transformed HR. Additive inverse of FEV_1_ (FEV_1_*-1) was used to allow better comparison of LCI with FEV_1_. *Definitions*: Crude model: unadjusted HR per one z-score increase in LCI and one z-score decrease in FEV_1_, Reduced model: Adjusted HR per one z-score increase in LCI and one z-score decrease in FEV_1_, adjusted for the selected variables sex, age, BMI, year of birth, number of hospitalisations; Final model: HR per one z-score increase in LCI and per one z-score decrease in FEV_1_, adjusted mutually in addition to the aforementioned variables. BMI = Body mass index, CF = Cystic fibrosis, LCI = Lung clearance index, LTX = Lung transplantation.

Effect sizes were influenced by the unequal variances of LCI and FEV_1_ in this CF population (Table 1 and 2). In the *Crude model* the HR for death or LTX was 1.30 [1.08 – 1.58] per 1.0 SD-score increase in LCI, and 1.55 [1.22 – 1.96] per 1.0 SD-score increase in FEV_1_. In the *Final model* the HR for death or LTX was 1.30 [1.06 – 1.60] per 1.0 SD-score increase in LCI, and 1.26 [0.91 – 1.74] per 1.0 SD-score decrease in FEV_1_.

Kaplan Meier survival curves show that individuals with LCI values above the population median LCI (7.3 z-score) had a higher risk of death or LTX compared to those with a LCI below the population median LCI at baseline (Figure 3). Findings for FEV_1_ were similar (Figure S2, OLS).

**Figure 3.**
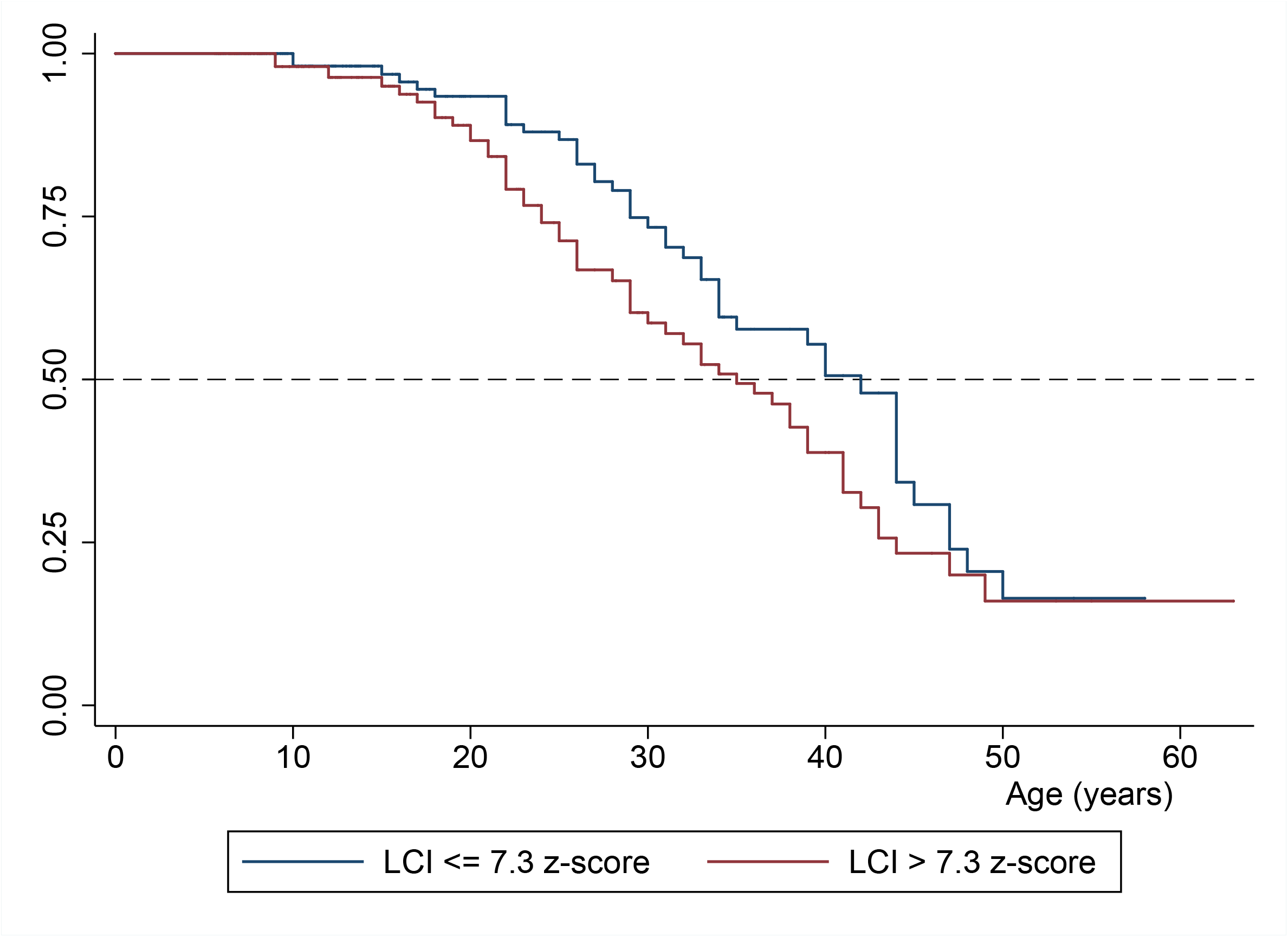
Respiratory survival in individuals with CF according to baseline LCI. Individuals with baseline LCI value ≤ study population median of 7.3 z-score, n = 119 are highlighted in blue. Individuals with baseline LCI value > study population median of 7.3 z-score, n = 118 are highlighted in red. *Definitions:* p = 0.50: 50% of the individuals in each group died or received LTX. BMI = Body mass index, CF = Cystic fibrosis, LCI = Lung clearance index.

**Figure 4.**
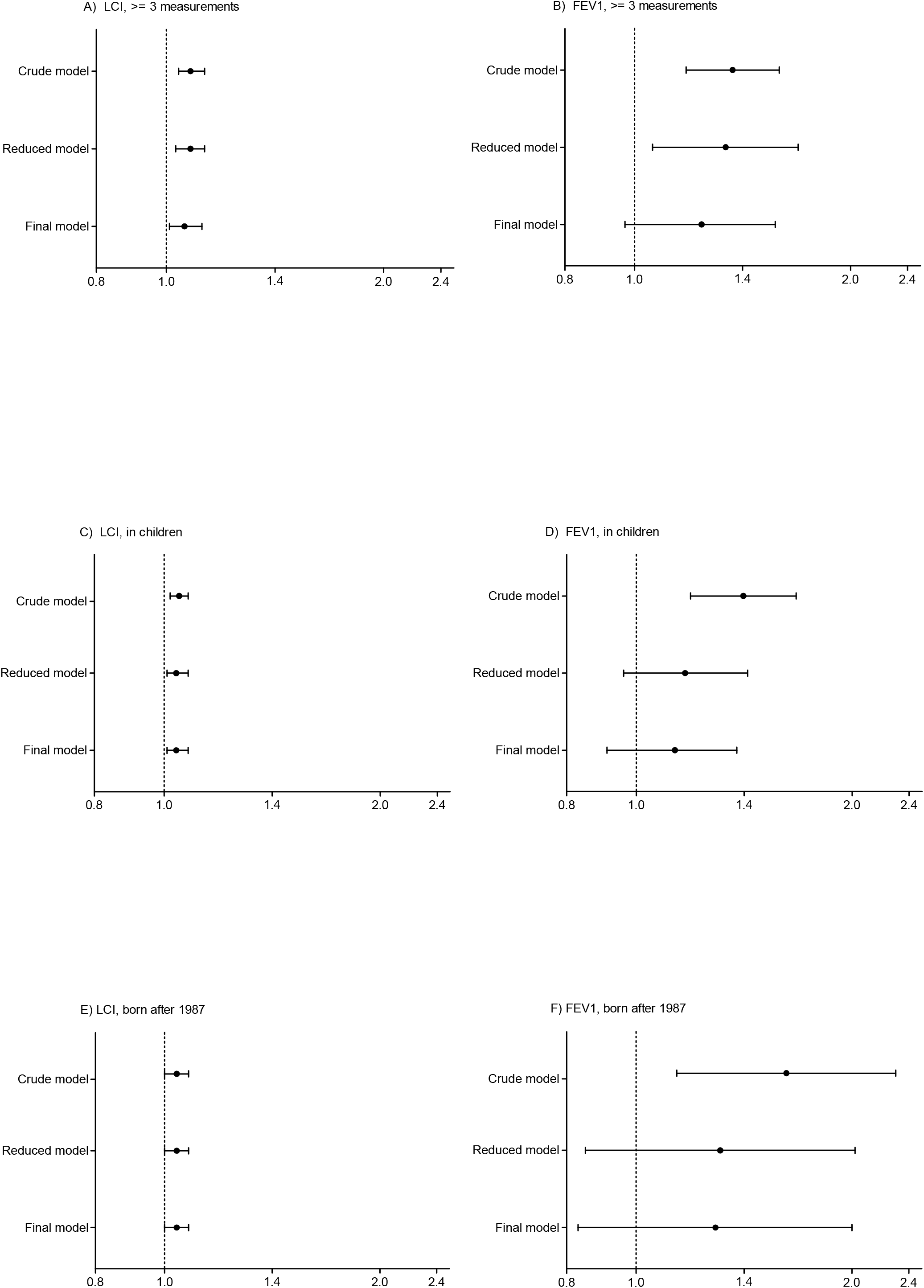
Risk of death of lung transplantation in the sensitivity analysis. Crude and adjusted Hazard Ratios [95% CI] for the risk of death or lung transplantation in the sensitivity analysis: *(i)* per one z-score increase in LCI A) and per one z-score decrease in FEV_1_ B) in individuals ≥ 3 LCI measurements within three years after study entry using the average of the first three available LCI and corresponding FEV_1_ values as baseline (n = 188), *(ii)* per one z-score increase in LCI C) and per one z-score decrease in FEV_1_ D) in children ≤ 16.0 years of age (n = 168) using the first available LCI and corresponding FEV_1_ value as baseline, and *(iii)* per one z-score increase in LCI E) and per one z-score increase in FEV_1_ F) in individuals born after 1987 (n = 102) using the first available LCI and corresponding FEV_1_ value as baseline . Additive inverse of FEV_1_ (FEV_1_*-1) was used to allow better comparison between LCI and FEV_1_. *Definitions*: Crude model: unadjusted HR per one z-score increase in LCI and one z-score decrease in FEV_1_, Reduced model: Adjusted HR per one z-score increase in LCI and one z-score decrease in FEV_1_, adjusted for the selected variables sex, age, BMI, year of birth, number of hospitalisations; Final model: HR per one z-score increase in LCI and per one z-score decrease in FEV_1_, adjusted mutually in addition to the aforementioned variables. BMI = Body mass index, CF = Cystic fibrosis, LCI = Lung clearance index, LTX = Lung transplantation.

### Sensitivity analyses

Sensitivity analyses were performed in subgroups of individuals, who (i) had three or more MBW tests within the first three years (n = 188, Table S5, OLS) of which the average LCI of the first three visits was derived as alternate baseline, or were (ii) born after 1987 (n = 102, Table 4), or (iii) aged ≤ 16.0 years (n = 168, Table 4), or (iv) had FEV_1_ ≥ -1.96 z-score (n = 108, Table S5, OLS). Sensitivity analyses confirmed the primary analysis: LCI predicted death or LTX in children aged ≤ 16.0 years and in younger individuals born after 1987. Using the mean over three LCI and FEV_1_ measurements as baseline values resulted in even higher estimates. The population characteristics in the sensitivity analyses are summarized in Table 3 and Table S4, OLS. Kaplan Meier survival curves are shown in Figure S3, OLS.

**Table 3.**
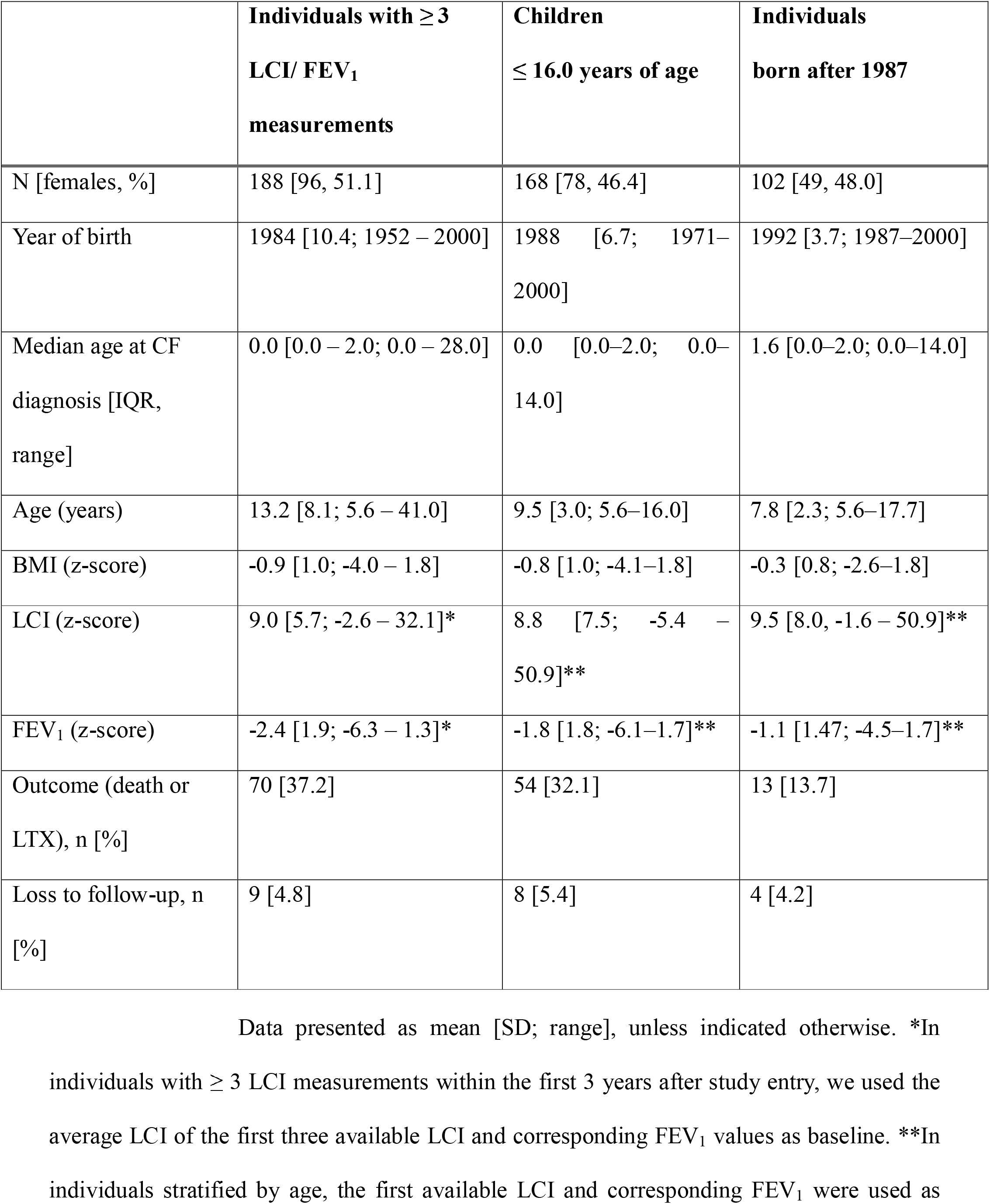
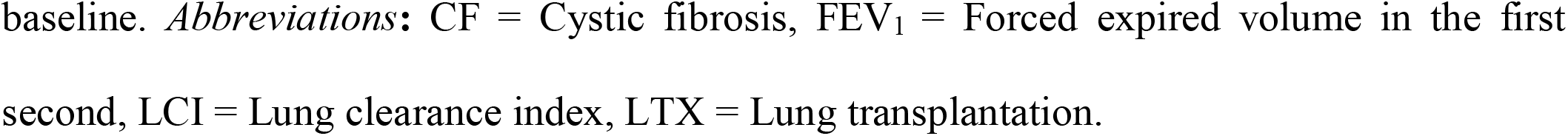
Population characteristics in the sensitivity analyses.

|  | **Individuals with $\geq 3$**<br>**LCI/ FEV<sub>1</sub>**<br>**measurements** | **Children**<br>**$\leq 16.0$ years of age** | **Individuals**<br>**born after 1987** |
| --- | --- | --- | --- |
| N [females, %] | 188 [96, 51.1] | 168 [78, 46.4] | 102 [49, 48.0] |
| Year of birth | 1984 [10.4; 1952 – 2000] | 1988 [6.7; 1971–<br>2000] | 1992 [3.7; 1987–2000] |
| Median age at CF<br>diagnosis [IQR,<br>range] | 0.0 [0.0 – 2.0; 0.0 – 28.0] | 0.0 [0.0–2.0; 0.0–<br>14.0] | 1.6 [0.0–2.0; 0.0–14.0] |
| Age (years) | 13.2 [8.1; 5.6 – 41.0] | 9.5 [3.0; 5.6–16.0] | 7.8 [2.3; 5.6–17.7] |
| BMI (z-score) | -0.9 [1.0; -4.0 – 1.8] | -0.8 [1.0; -4.1–1.8] | -0.3 [0.8; -2.6–1.8] |
| LCI (z-score) | 9.0 [5.7; -2.6 – 32.1]* | 8.8 [7.5; -5.4 –<br>50.9]** | 9.5 [8.0, -1.6 – 50.9]** |
| FEV<sub>1</sub> (z-score) | -2.4 [1.9; -6.3 – 1.3]* | -1.8 [1.8; -6.1–1.7]** | -1.1 [1.47; -4.5–1.7]** |
| Outcome (death or<br>LTX), n [%] | 70 [37.2] | 54 [32.1] | 13 [13.7] |
| Loss to follow-up, n<br>[%] | 9 [4.8] | 8 [5.4] | 4 [4.2] |

baseline. *Abbreviations:* CF = Cystic fibrosis, FEV<sub>1</sub> = Forced expired volume in the first second, LCI = Lung clearance index, LTX = Lung transplantation.

**Table 4.**
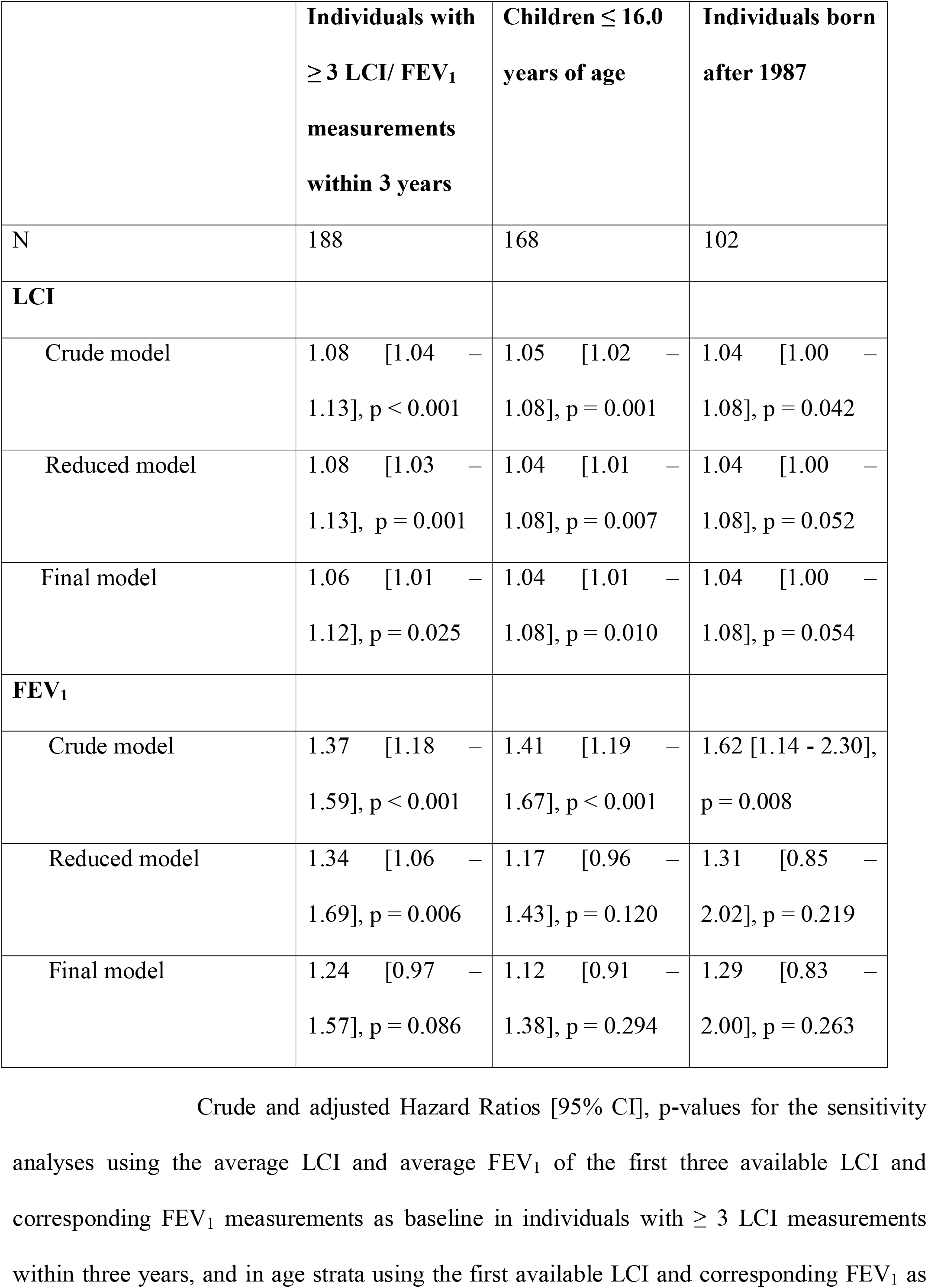
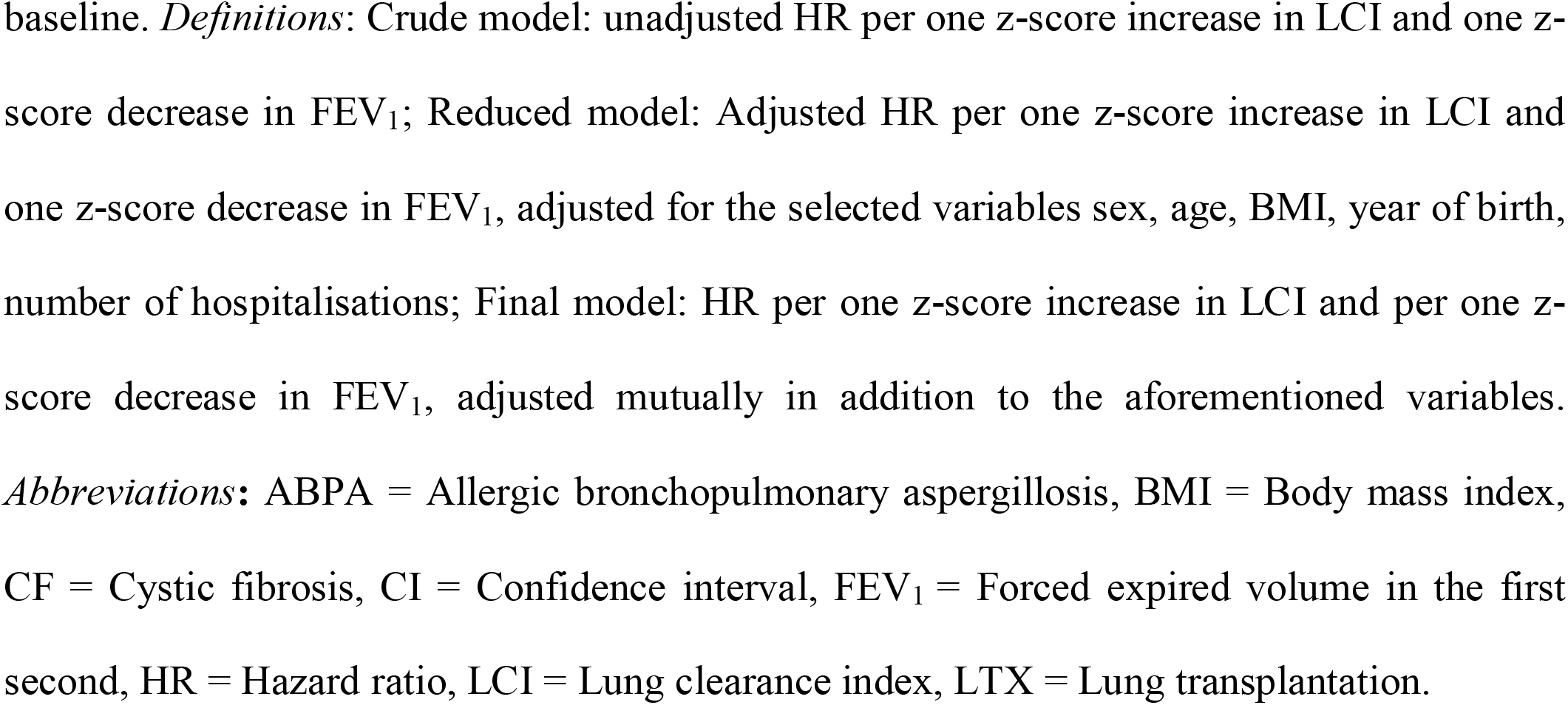
Risk of death or lung transplantation in the sensitivity analyses.

|  | **Individuals with**<br>**$\geq 3$ LCI/ FEV<sub>1</sub>**<br>**measurements**<br>**within 3 years** | **Children $\leq 16.0$**<br>**years of age** | **Individuals born**<br>**after 1987** |
| --- | --- | --- | --- |
| N | 188 | 168 | 102 |
| **LCI** |  |  |  |
| Crude model | 1.08 [1.04 – 1.13], p < 0.001 | 1.05 [1.02 – 1.08], p = 0.001 | 1.04 [1.00 – 1.08], p = 0.042 |
| Reduced model | 1.08 [1.03 – 1.13], p = 0.001 | 1.04 [1.01 – 1.08], p = 0.007 | 1.04 [1.00 – 1.08], p = 0.052 |
| Final model | 1.06 [1.01 – 1.12], p = 0.025 | 1.04 [1.01 – 1.08], p = 0.010 | 1.04 [1.00 – 1.08], p = 0.054 |
| **FEV<sub>1</sub>** |  |  |  |
| Crude model | 1.37 [1.18 – 1.59], p < 0.001 | 1.41 [1.19 – 1.67], p < 0.001 | 1.62 [1.14 - 2.30], p = 0.008 |
| Reduced model | 1.34 [1.06 – 1.69], p = 0.006 | 1.17 [0.96 – 1.43], p = 0.120 | 1.31 [0.85 – 2.02], p = 0.219 |
| Final model | 1.24 [0.97 – 1.57], p = 0.086 | 1.12 [0.91 – 1.38], p = 0.294 | 1.29 [0.83 – 2.00], p = 0.263 |

baseline. *Definitions:* Crude model: unadjusted HR per one z-score increase in LCI and one z-score decrease in FEV<sub>1</sub>; Reduced model: Adjusted HR per one z-score increase in LCI and one z-score decrease in FEV<sub>1</sub>, adjusted for the selected variables sex, age, BMI, year of birth, number of hospitalisations; Final model: HR per one z-score increase in LCI and per one z-score decrease in FEV<sub>1</sub>, adjusted mutually in addition to the aforementioned variables. *Abbreviations:* ABPA = Allergic bronchopulmonary aspergillosis, BMI = Body mass index, CF = Cystic fibrosis, CI = Confidence interval, FEV<sub>1</sub> = Forced expired volume in the first second, HR = Hazard ratio, LCI = Lung clearance index, LTX = Lung transplantation.

## Discussion

### Summary

This is the first study to show that LCI is associated with survival in individuals with CF. We found that per one z-score increase in LCI, the risk (HR) of death or LTX increased on average by 4%. After adjustment for heterogeneous variance of lung function values in the CF population, the risk of death or LTX increased on average by 30% per one SD-score increase in LCI. We verified this association in regression models adjusting for clinical and anthropometric variables and in sensitivity analyses. Baseline lung function values averaged across three visits provided stronger association with death or LTX compared to baseline lung function including single LCI and FEV_1_ values only. LCI predicted death or LTX in children and younger individuals (born after 1987).

### Association of lung function with respiratory disease outcomes

This study links LCI with respiratory survival in individuals with CF. These data are essential for biomarkers to be recognized as novel surrogate end-points of long-term prognosis[8]. LCI was associated with death or LTX after adjusting for known risk factors [26]. As expected, several variables were removed from the final model, as they either were strongly correlated or did not contribute to the prediction of survival. Few other studies assessed the degree to which LCI may predict the clinical course in CF. In school-aged children with CF, the risk of future pulmonary exacerbations increased by 12% after an LCI increase of [0.5 units at baseline[27]. We have recently demonstrated that per one LCI unit increase, the risk of future pulmonary exacerbation increased by 13% in children and adults with Primary Ciliary Dyskinesia[28]. These data suggest that LCI is predictive of adverse respiratory events in muco-obstructive lung diseases.

We confirmed that FEV_1_ continues to be an important surrogate endpoint of survival in CF[3, 29]. However, after adjusting FEV_1_ with LCI, the association between FEV_1_ and survival was weakened substantially. In line with previous findings, the variables sex, age, BMI and number of hospitalisations partially explained the association of FEV_1_ with survival[26]. We assume that the ability of FEV_1_ to predict survival may further decline in the current era of CF care including CFTR modulators and evolving survival patterns[1, 26]. There are several reasons why LCI should be further considered. FEV_1_ may remain normal in childhood and studies assessing treatment regimens targeted to improve lung function may be constrained by ceiling effects from normal FEV_1_[30]. In addition, the predictive value of FEV_1_ for structural lung disease is poor. LCI is a reliable measure of global ventilation inhomogeneity arising from central and mainly peripheral airways[31]. MBW is characterized by a high feasibility and good repeatability[32, 33]. We and others have demonstrated the validity of LCI, *i*.*e*. the correlation with ‘‘gold standard’’ outcomes such as infection with *Pseudomonas aeruginosa* and structural lung disease[4, 6].

We showed that both, a single “snap shot” LCI measurement and averaged triplicate LCI measurements predicted respiratory survival. LCI averaged across three visits provided even stronger association with death or LTX compared to a single LCI value. Clinically important events such as ABPA or pulmonary exacerbations contribute to LCI variability and may therefore influence the predictive capacity of LCI [34, 35]. In our study, LCI, but not FEV_1_, was predictive of death or LTX in younger individuals and individuals with mild CF lung disease. LCI seems to deteriorate more rapidly compared to FEV_1_[10, 26]. We assume that clinical care improved during the study and possibly altered lung disease phenotypes from multi-airway generations obstruction in older individuals to mainly peripheral airway generations obstruction in younger individuals[9]. The influence of birth year on the association of FEV_1_ with survival was more pronounced compared to LCI, which indicated a temporal trend in our study. We considered birth year, age at CF diagnosis and age at baseline as proxies for improvements in clinical care and decreasing annual death rate in CF and adjusted for the latter variables.

### Strengths and limitations

Major strengths of our study are the large sample size and wide spectrum of disease severity including individuals born over five decades, and the low number of missing values. While prevalence of *Pseudomonas aeruginosa* was greater compared to most contemporary cohorts, spirometry indices were comparable. We acknowledge the historical treatment regimens in our study. As the clinical course usually differs between pre- and post-LTX, we did not include deaths post-LTX as study outcome[26]. Censoring subjects at the date of LTX may have slightly underestimated the person-years at risk. Further studies are required to externally validate our findings in large populations. Yet, current CF cohorts investigating LCI started 15 years ago and therefore the association with survival can be studied earliest in one or two decades[36, 37].

The CF centre Bern already had extensive experience in MBW at the time of our study and has collated one of the largest LCI datasets to our knowledge[10, 19]. N_2_MBW was performed regularly during clinical visits but did not influence clinical decisions, reducing the risk of selection bias or confounding by indication. The N_2_MBW setup was standard at that time and remained unchanged during the study, but is no longer available. It appears unlikely that measurement error would have positively confounded the association of LCI with survival. Our findings appear relatively independent of evolving MBW technologies as suggested by the consistent risk estimates derived from normalized LCI values calculated from an external healthy population and the current study population. Thereby we also accounted for unequal variance of lung function indices in the study population using parametric (SD-score based on the study population) and non-parametric (median LCI based on the study population) approaches. Yet, absolute LCI values in our study should be interpreted cautiously. The upper limit of normal LCI was 9.5 units which is higher compared to current N_2_MBW setups[19, 38].

### Clinical considerations

Our data support the use of LCI in routine clinical surveillance of individuals with CF. Time to death or LTX in individuals with higher LCI values was shorter compared to those with lower LCI. Future decision algorithms to recommend LTX may need to include LCI. LCI is increasingly used as study endpoint of trials assessing efficacy of CFTR modulators in children[39]. Recent evidence suggests that LCI correlates with the extent of naïve CFTR function suggesting LCI as a candidate treatable trait[40]. Increase in LCI could be used to timely initiate CFTR modulator treatment in otherwise asymptomatic individuals with CF and normal FEV_1_.

### Conclusion

The association between LCI and death or LTX may be considered as a milestone in the establishment of LCI as surrogate endpoint of survival. By measuring LCI, health care providers and individuals with CF may gain better understanding of possible implications of increments in ventilation inhomogeneity. Further research is necessary to determine whether LCI guided treatment improves respiratory disease outcomes.

## Data Availability

The data that support the findings of this study are available from the corresponding author, Florian Singer, upon reasonable request.

## Acknowledgements

The authors thank to all individuals and families for allowing their MBW and clinical data to be used for research. The authors thank Christian Benden^1^, Carmen Casaulta^2^, Gianluca Calderari^3^, Kathleen Jahn^4^, Angela Koutsokera^5^Anna Ruedeberg^2^, Bernhard Schwizer^6^, Alain Sauty^7^, Michael Tamm^8^, Gisela Wirz^2^, Maura Zanolari^3,9^, and all other collaborators helping with data collection.

^1^Division of Pulmonary Medicine, University Hospital of Zurich, Zurich, Switzerland

^2^Division of Respiratory Medicine, Department of Paediatrics, Inselspital University Hospital Bern, University of Bern, Bern, Freiburgstrasse 15, 3010 Bern, Switzerland

^3^Studio Medico, Corso Pestalozzi 11, 6900 Lugano, Switzerland

^4^Department of Pneumology, University Hospital Basel, Switzerland

^5^Service de pneumologie, centre hospitalier universitaire Vaudois, 1011 Lausanne, Suisse

^6^Department of Nuclear Medicine and Radiology, Cantonal Hospital Lucerne, Lucerne, Switzerland

^7^Pneumologist, Service de Pneumologie, Hôpital Neuchâtelois, Neuchâtel, Switzerland

^8^Dept of Internal Medicine, University Hospital, Basel, Switzerland

^9^Dept of Paediatrics, Hospital of Bellinzona, Bellinzona, Switzerland

---

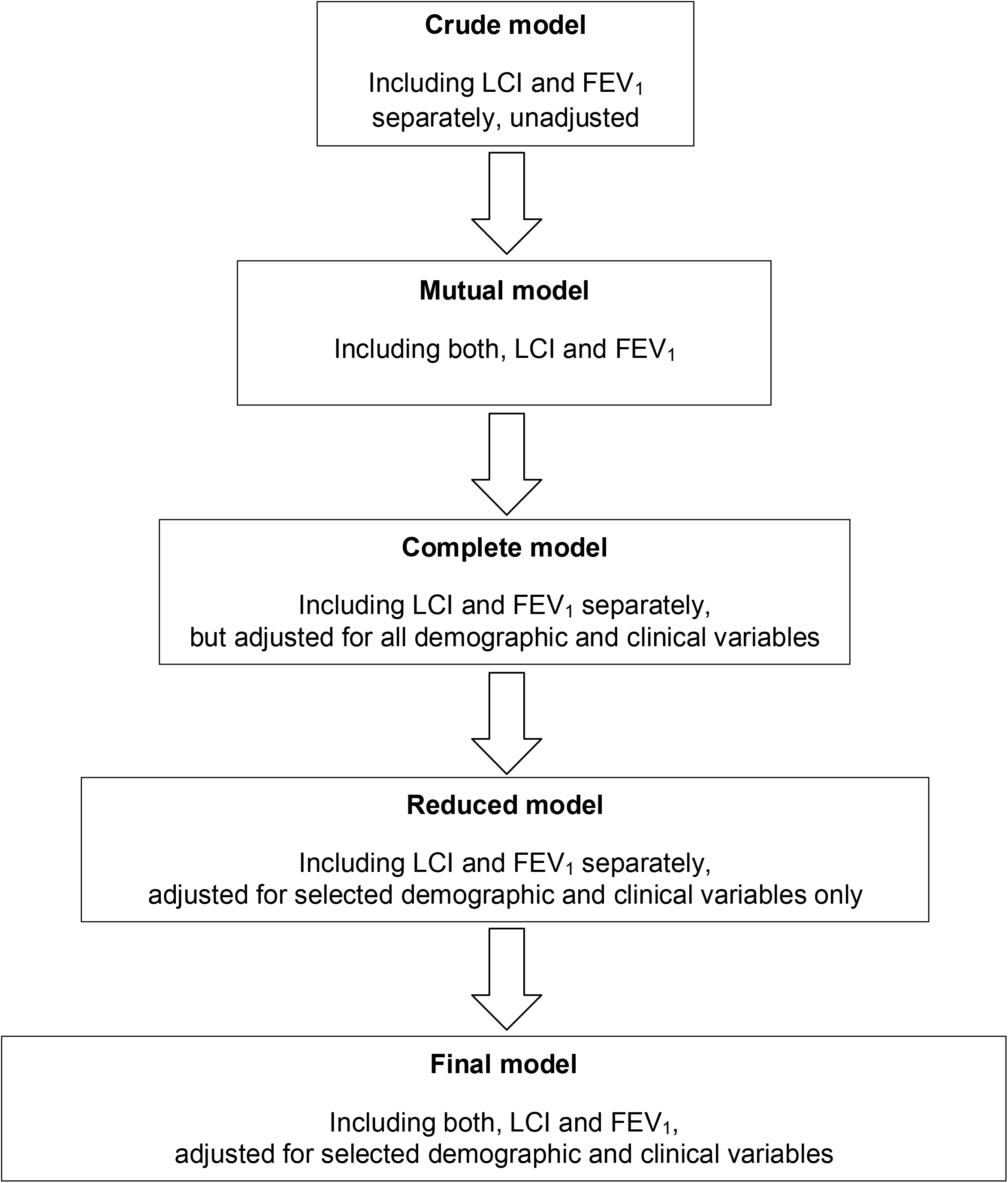

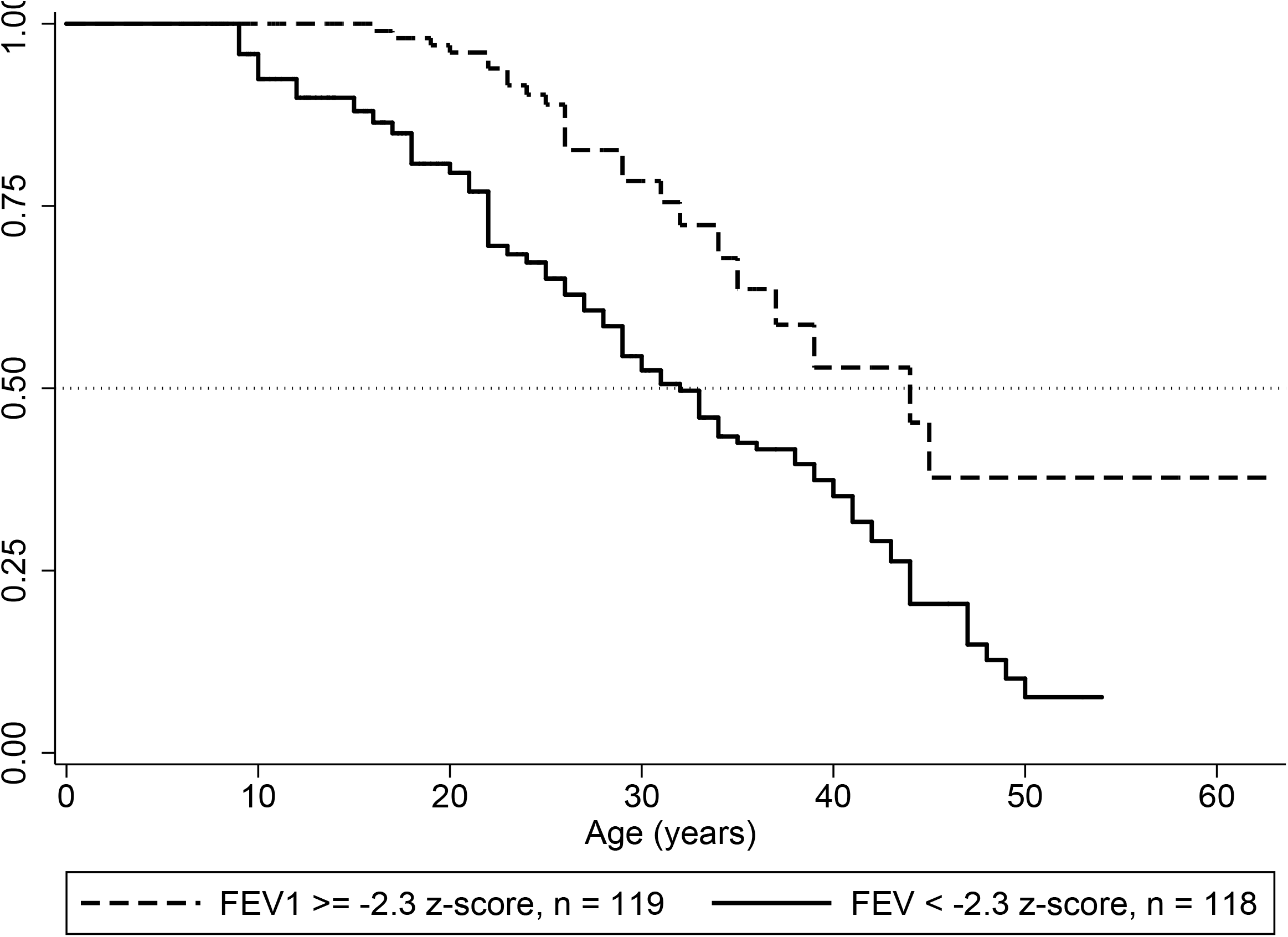

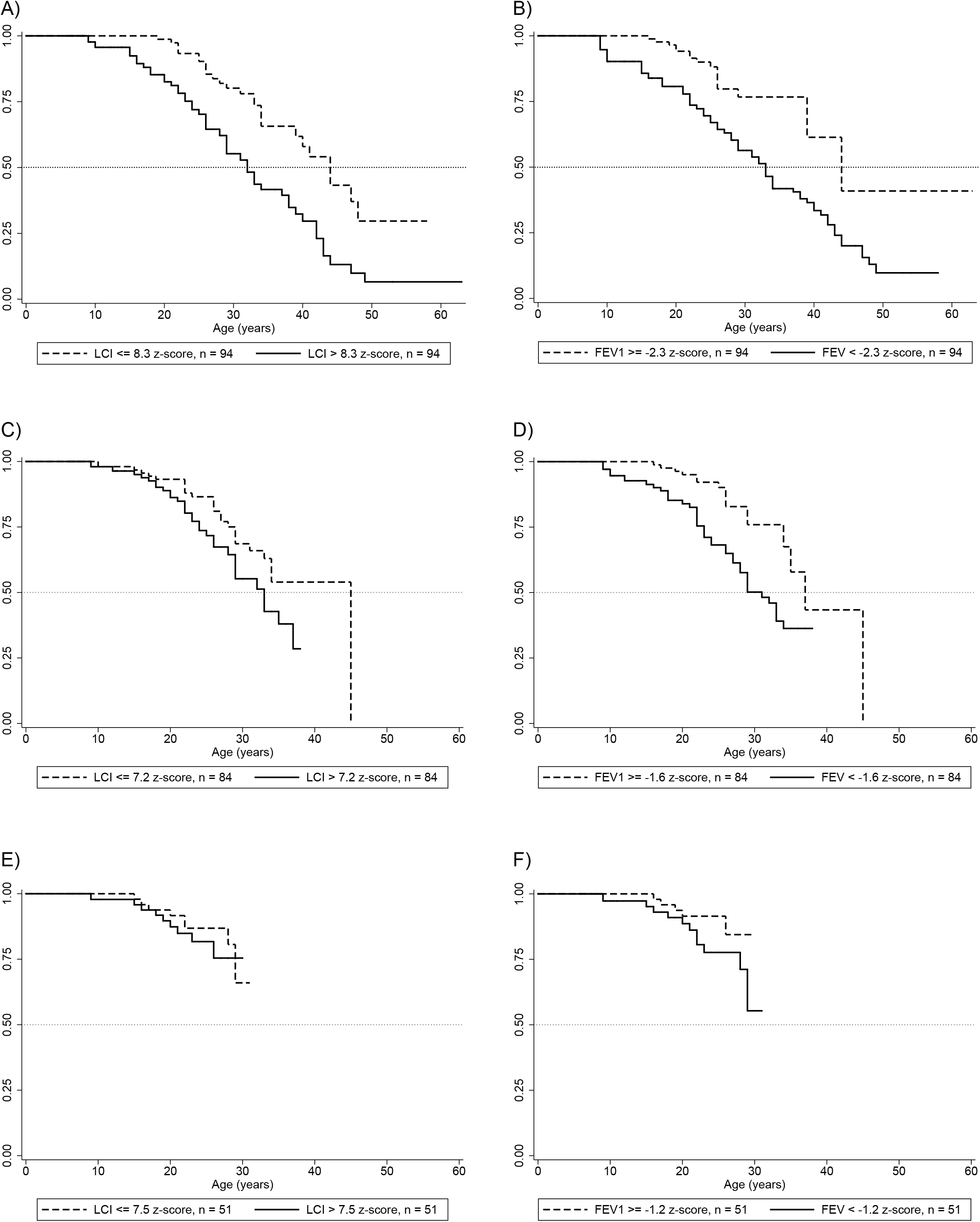

