## Supplement for "Association of lung clearance index with survival in individuals with cystic fibrosis"

**Tables**

**Table S1: Definitions of variables in baseline and sensitivity analyses.**

| **Sex** | Gender, binary (0: female, 1: male) |
| --- | --- |
| **Age at CF diagnosis** | Individuals age at CF diagnosis in years |
| **Mutation** | Individuals homozygote for F508del (0)*, individuals heterozygote for F508del (1),*  individuals with any other mutation (2) |
| **Test date** | Date (yyyy/mm/dd) of the third LCI measurement out of the three LCI measurements within the first three years after study entry; begin of crude mortality time |
| **Age, height, weight, BMI** | Values from test date (age: years, height: meters, weight: kg, BMI: z-score) |
| **LCI** | *Baseline*: First available LCI measurement as baseline value  *Sensitivity analysis:* Mean of the first three LCI values within the first three years after study entry (= first available LCI measurement) as baseline value and exclusion of individuals with less than three measurements within the first three years after study entry |
| **FEV_1_** | *Baseline*: FEV_1_ value that was reported corresponding to the first available LCI value (FEV_1_ and LCI were measured at the same day)  Sensitivity analysis: Mean of the three FEV_1_ values that were reported corresponding to the first three available LCI values within the first three years after study entry |
| **Hospitalisations** | Count of all hospitalisations reported in the electronic patient charts within the first three years after study entry |
| **Exacerbations** | Count of all exacerbations reported in the electronic patient charts within the first three years after study entry |
| **Pancreatic insufficiency** | Pancreatic insufficiency at least once reported/ diagnosed within the frist three years after study entry “yes – 1”, otherwise “no - 0” |
| **CFRD** | CFRD at least once reported/ diagnosed within the first three years after study entry “yes – 1”, otherwise “no - 0” |
| **ABPA** | ABPA at least once reported/ diagnosed within the first three years after study entry “yes – 1”, otherwise “no - 0” |
| **Microbiology** | Pseudomonas aeruginosa at least once reported/ diagnosed within the first three years after study entry “yes – 1”, otherwise “no – 0” |
|  | Staphylococcus aureus at least once reported/ diagnosed within the first three years afters study entry “yes – 1”, otherwise “no – 0” |
| **Medication** | At least once treated with any antibiotics (oral, iv, inhaled) within the first three years after study entry “yes – 1”, otherwise “no – 0” |
|  | At least once treated with any inhaled medication (mucolytics, bronchodilators, steroids) within the first three years after study entry “yes – 1”, otherwise “no – 0” |
| **Outcome (death or LTX)** | Date (yyyy/mm/dd) of death or of lung transplantation as reported in the electronic patient charts (1) versus survival (0). Survival means the patient was alive until the end of the study in 2018/12/31, e.g. when the patient had at least one follow-up visit after 2018/12/31) |

**Legend Table S1:** *Abbreviations***:** ABPA = Allergic bronchopulmonary aspergillosis, BMI = Body mass index, CF = Cystic fibrosis, CFRD = Cystic fibrosis-related diabetes, FEV_1_ = Forced expired volume in the first second, LCI = Lung clearance index, LTX = Lung transplantation

**Table S2:** **Population characteristics of individuals stratified by endpoint.**

|  | **Death** | **LTX** | **Alive** | **Lost to follow-up** |
| --- | --- | --- | --- | --- |
| N [females, %] | 41 [22, 53.7] | 53 [27, 50.9] | 128 [54, 42.2] | 15 [10, 66.7] |
| LCI (units) | 17.2 [7.3; 3.9 – 46.6] | 18.9 [8.4; 5.6 – 59.8] | 16.9 [6.6; 3.5 – 47.2] | 19.0 [9.2; 9.3 – 41.2] |
| LCI (z-score) | 8.3 [7.3; - 5.0 – 37.7] | 10.0 [8.4; -3.3 – 50.9] | 8.0 [6.6; -5.4 – 38.3] | 10.1 [9.2; 0.4 – 32.3] |
| FEV_1_ (z-score) | -3.7 [1.9; -6.3 – 0.8] | -3.1 [1.8; -6.1 – 0.4] | -1.6 [1.6; -5.6 – 1.7] | -3.5 [1.9; -6.4 – -0.5] |
| Year of birth | 1974 [9.4; 1952 – 1991] | 1980 [8.7; 1960 – 1997] | 1988 [9.8; 1955 – 2000] | 1980 [7.7; 1963 – 1990] |
| Age at baseline (years) | 19.7 [9.7; 6.0 – 41.0] | 15.0 [7.4; 5.9 – 34.2] | 11.4 [7.1; 5.6 – 40.2] | 15.8 [6.4; 6.9 – 26.1] |
| BMI (z-score) | -1.3 [1.2; -4.0 – 0.3] | -1.2 [1.0; -4.0 – 1.4] | -0.5 [1.0; -3.4 – 2.4] | -1.4 [1.1; -4.0 – 0.0] |
| Age at outcome (years) | 32.0 [10.8; 9.0 – 50.0] | 28.8 [9.0; 15.0 – 47.0] | 30.4 [9.8; 18.0 – 63.0] | 25.3 [11.4; 8.0 – 44.0] |

**Legend Table S2:** Data presented as mean [SD; range], unless indicated otherwise. *Abbreviations:* BMI = Body mass index, FEV_1_ = Forced expired volume in the first second, LCI = Lung clearance index.

**Table S3. Risk of death or lung transplantation according to baseline lung function.**

|  | **LCI** | **FEV_1_** |
| --- | --- | --- |
| Crude model | 1.04 [1.01 – 1.06] | 1.25 [1.11 – 1.41], |
| Mutual model | 1.03 [1.00 – 1.06] | 1.22 [1.08 – 1.38] |
| Complete model | 1.03 [1.00 – 1.06] | 1.15 [0.97 – 1.36] |
| Reduced model | 1.04 [1.01 – 1.07] | 1.18 [1.01 – 1.38] |
| Final model | 1.04 [1.01 – 1.07] | 1.12 [0.95 – 1.33], |

**Legend Table S3:** Crude and adjusted Hazard Ratios [95% CI] for the risk of death or lung transplantation using the first available LCI and corresponding FEV_1_ value as baseline. *Definitions:* Crude model: unadjusted HR per one z-score increase in LCI and one z-score decrease in FEV_1_; Mutual model: HR per one z-score increase in LCI and one z-score decrease in FEV_1_, adjusted mutually; Complete model: HR per one z-score increase in LCI and per one z-score decrease in FEV_1_ adjusted separately for all demographic and clinical variables (sex, age, BMI, year of birth, number of hospitalisations, number of exacerbations, mutation, pancreatic insufficiency, CFRD, ABPA, microbiology, medication), Reduced model: Adjusted HR per one z-score increase in LCI and one z-score decrease in FEV_1_, adjusted for the selected variables sex, age, BMI, year of birth, number of hospitalisations; Final model: HR per one z-score increase in LCI and per one z-score decrease in FEV_1_, adjusted mutually in addition to the aforementioned variables. *Abbreviations*: ABPA = Allergic bronchopulmonary aspergillosis, BMI = Body mass index, CF = Cystic fibrosis, CFRD = Cystic fibrosis-related diabetes, CI = Confidence interval, FEV_1_ = Forced expired volume in the first second, HR = Hazard ratio, LCI = Lung clearance index, LTX = Lung transplantation.

**Table S4: Population characteristics in individuals with normal FEV_1_ and in adults.**

|  | **Individuals with normal FE**V_1_ **(≥ - 1.96 z-score)** | **Adults**  **(≥ 16.0 years of age)** |
| --- | --- | --- |
| N [females, %] | 108 [55, 50.9] | 69 [35, 50.7] |
| Year of birth | 1988 [9.2; 1955 – 2000] | 1970 [6.8; 1952–1988] |
| Median age at CF diagnosis [IQR, range] | 1.0 [0.0 – 2.0; 0.0 – 14.0] | 1.0 [0.0–3.0; 0.0–28.0] |
| Age (years) | 10.4 [6.2; 5.6 – 40.2] | 24.8 [6.5; 16.1–41.0] |
| BMI at prediction start (z-score) | -0.4 [1.0; -4.2 – 2.4] | -1.1 [1.2; -4.2–0.4] |
| LCI (z-score) | 7.3 [7.5; -5.4 – 50.9] | 8.3 [6.8; -5.0 – 32.3] |
| FEV_1_ ( z-score) | -0.56 [0.9; -1.9 – 1.7] | -3.8 [1.8; -6.4–0.4] |
| Outcome (Death or LTX), n [%] | 24 [22.2] | 40 [58.0] |
| Loss to follow-up, n [%] | 4 [3.7] | 7 [10.14] |

**Legend Table S4:** Data presented as mean [SD, range], unless indicated otherwise. *Definitions:* Baseline = First available LCI and corresponding FEV_1_ value demographic and clinical data derived within the first three years after study entry; *Abbreviations***:** BMI = Body mass index, CF = Cystic fibrosis, FEV_1_ = Forced expired volume in the first second, LCI = Lung clearance index, LTX = Lung transplantation.

**Table S5: Risk of death or lung transplantation individuals with normal FEV_1_ and in adults.**

|  | **Individuals with normal FEV_1_ (≥ - 1.96 z-score)** | **Adults**  **(> 16.0 years of age)** |
| --- | --- | --- |
| N | 108 | 69 |
| **LCI** |  |  |
| Crude model | 1.02 [0.97 – 1.08] | 1.01 [0.96 – 1.06] |
| Mutual model | 1.03 [0.98 – 1.08] | 0.98 [0.92 – 1.03] |
| Complete model | 1.01 [0.96 – 1.07] | 1.01 [0.94 – 1.09] |
| Reduced model | 1.02 [0.97 – 1.07] | 1.00 [0.95 – 1.06] |
| Final model | 1.03 [0.98 – 1.09] | 0.97 [0.91 – 1.04] |
| **FEV_1_** |  |  |
| Crude model | 0.99 [0.61 – 1.61] | 1.33 [1.07 – 1.65] |
| Mutual model | 0.96 [0.58 – 1.57] | 1.37 [1.09 – 1.72] |
| Complete model | 0.68 [0.35 – 1.31] | 1.22 [0.92 – 1.62] |
| Reduced model | 0.76 [0.43 – 1.36] | 1.26 [0.96 – 1.64] |
| Final model | 0.71 [0.40 – 1.29] | 1.33 [0.99 – 1.79] |

**Legend Table S5:** Crude and adjusted Hazard Ratios [95% CI] for the risk of death or lung transplantation in individuals with normal FEV_1_ (≥ - 1.96 z-score) and adults (≥ 16.0 years of age) using the first available LCI and corresponding FEV_1_ value as baseline. *Definitions:* Crude model: unadjusted HR per one z-score increase in LCI and one z-score decrease in FEV_1_; Mutual model: HR per one z-score increase in LCI and one z-score decrease in FEV_1_, adjusted mutually; Complete model: HR per one z-score increase in LCI and per one z-score decrease in FEV_1_ adjusted separately for all demographic and clinical variables (sex, age, BMI, year of birth, number of hospitalisations, number of exacerbations, mutation, pancreatic insufficiency, CFRD, ABPA, microbiology, medication), Reduced model: Adjusted HR per one z-score increase in LCI and one z-score decrease in FEV_1_, adjusted for the selected variables sex, age, BMI, year of birth, number of hospitalisations; Final model: HR per one z-score increase in LCI and per one z-score decrease in FEV_1_, adjusted mutually in addition to the aforementioned variables. *Abbreviations***:** ABPA = Allergic bronchopulmonary aspergillosis, BMI = Body mass index, CF = Cystic fibrosis, CFRD = Cystic fibrosis-related diabetes, CI = Confidence interval, FEV_1_ = Forced expired volume in the first second, HR = Hazard ratio, LCI = Lung clearance index, LTX = Lung transplantation.

**Table S6: Fully adjusted Cox proportional hazards regression model.**

|  | **HR** | **P** | **[95% CI]** |
| --- | --- | --- | --- |
| LCI (z-score) | 1.033 | 0.030 | 1.003 – 1.064 |
| FEV_1_ (z-score) | 1.099 | 0.294 | 0.921 – 1.310 |
| Sex (0 = female, 1 = male) | 0.486 | 0.003 | 0.302 – 0.783 |
| Age (years) | 0.892 | 0.002 | 0.830 – 0.958 |
| BMI (z-score) | 0.771 | 0.034 | 0.606 – 0.980 |
| Year of birth | 0.947 | 0.104 | 0.886 – 1.011 |
| Age at CF-diagnosis | 0.998 | 0.952 | 0.936 – 1.064 |
| Mutation (0 = F508del homozygous, 1 = F508del heterozygous, 2 = Other) | 1.027 | 0.910 | 0.652 – 1.615 |
| Number of exacerbations | 1.075 | 0.277 | 0.944 – 1.224 |
| Number of hospitalisations | 1.129 | 0.196 | 0.939 – 1.357 |
| ABPA (1= yes, 0 = no) | 1.595 | 0.238 | 0.734 – 3.466 |
| Pseudomonas aeruginosa (1= yes, 0 = no) | 1.148 | 0.637 | 0.606 – 2.176 |
| Staphylococcus aureus (1= yes, 0 = no) | 0.679 | 0.122 | 0.415 – 1.109 |
| CFRD (1= yes, 0 = no) | 0.873 | 0.693 | 0.444 – 1.715 |
| Pancreas insufficiency (1= yes, 0 = no) | 0.895 | 0.866 | 0.245 – 3.272 |
| Antibiotic medication (1 = yes, 0 = no) | 1.079 | 0.911 | 0.287 – 4.058 |

**Legend Table S6:** Estimates for the fully adjusted Cox proportional hazard regression model using the first available LCI and corresponding FEV_1_ value as baseline. *Abbreviations:* ABPA = Allergic bronchopulmonary aspergillosis, BMI = Body mass index, CF = Cystic fibrosis, CFRD = CF-related diabetes, CI = Confidence interval, FEV_1_ = Forced expired volume in the first second, HR = Hazard ratio, LCI = Lung clearance index, P = P-value.

**Table S7: Final Cox proportional hazards regression model.**

|  | **HR** | **P** | **[95% CI]** |
| --- | --- | --- | --- |
| LCI (z-score) | 1.037 | 0.011 | 1.008 – 1.066 |
| FEV_1_ (z-score) | 1.124 | 0.164 | 0.953 – 1.327 |
| Sex (0 = female, 1 = male) | 0.570 | 0.011 | 0.370 – 0.878 |
| Age (years) | 0.907 | 0.005 | 0.847 – 0.971 |
| BMI (z-score) | 0.772 | 0.027 | 0.613 – 0.971 |
| Year of birth | 0.952 | 0.109 | 0.896 - 1.011 |
| Number of hospitalisations | 1.117 | 0.151 | 0.961 – 1.300 |

**Legend Table S7:** Estimates for the final Cox proportional hazards regression model using the first available LCI and corresponding FEV_1_ value as baseline. Definition: Final model. HR per one z-score increase in LCI and one z-score decrease in FEV_1_ adjusted mutually in addition to the selected variables (sex, age, BMI, birth year, number of hospitalisations). *Abbreviations:* BMI = Body mass index, CI = Confidence interval, FEV_1_ = Forced expired volume in the first second, HR = Hazard ratio, LCI = Lung clearance index, P = P-Value

**Figure legends**

**Figure S1. Analysis steps.** *Crude model:* Unadjusted HRs per z-score increase in LCI and decrease in FEV_1_. *Mutual model:* HR per z-score increase in LCI and decrease in FEV_1_, adjusted mutually; *Complete model:* HR per z-score increase in LCI and decrease in FEV_1_, adjusted separately for all demographic and clinical variables (sex, age, BMI, year of birth, mutation, age at CF diagnosis, infection burden, CFRD, ABPA, pancreas function, medication, number of exacerbations and hospitalisations); *Reduced model:* HR per z-score increase in LCI and decrease in FEV_1_, adjusted separately for selected variables only (sex, age, BMI, year of birth, number of hospitalisations); *Final model:* HR per z-score increase in LCI and decrease in FEV_1_, adjusted mutually in addition to selected variables (sex, age, BMI, year of birth, number of hospitalisations). ABPA = Allergic bronchopulmonary aspergillosis, BMI = Body mass index, CF = Cystic fibrosis, CFRD = Cystic fibrosis-related diabetes, FEV_1_ = Forced expired volume in the first second, HR = Hazard ratio, LCI = Lung clearance index.

**Figure S2. Respiratory survival in individuals with CF according to baseline FEV_1_.** Individuals with baseline FEV_1_ values ≥ study population median of -2.3 z-score, n = 119 (dashed line) vs. individuals with baseline FEV_1_ values < study population median of -2.3 z-score, n = 118 (solid line) using the first available FEV_1_ value as baseline. CF = Cystic fibrosis, FEV_1_ = Forced expired volume in the first second, LTX = Lung transplantation, p = 0.50: 50% of the individuals in each group died or received LTX.

**Figure S3. Respiratory survival in the sensitivity analyses.** *A)* Individuals with ≥ 3 LCI measurements within 3 years and average baseline LCI values ≤ study population median of 8.3 z-score, n = 84 (dashed line) vs. individuals with average baseline LCI values > study population median of 8.3 z-score, n = 84 (solid line). *B)* Individuals with ≥ 3 LCI measurements within 3 years and average baseline FEV_1_ values ≥ study population median of -2.3 z-score, n = 94 (dashed line) vs. adults with average baseline FEV_1_ values < study population median of -2.3 z-score, n = 94 (solid line); *C)* Children (≤ 16.0 years of age) with baseline LCI value ≤ study population median of 7.2 z-score, n = 84 (dashed line) vs. children with baseline LCI value > study population median of 7.2 z-score, n = 84 (solid line); *D)* Children (≤ 16.0 years of age) with baseline FEV_1_ value ≥ study population median of -1.6 z-score, n = 84 (dashed line) vs. children with baseline FEV_1_ value < study population median of -1.6 z-score, n = 84 (solid line). *E)* Individuals born after 1987 with baseline LCI value ≤ study population median of 7.5 z-score, n = 51 (dashed line) vs. individuals with baseline LCI value > study population median of 7.5 z-score, n = 51 (solid line). *F)* Individuals born after 1987 with baseline FEV_1_ value ≥ study population median of -1.2 z-score, n = 51 (dashed line) vs. individuals with baseline FEV_1_ value < study population median of -1.2 z-score, n = 51 (solid line). CF = Cystic fibrosis, FEV_1_ = Forced expired volume in the first second, LCI = Lung clearance index, LTX = Lung transplantation, p = 0.50: 50% of the individuals in each group died or received LTX.
